# What can data mining tell us about patient safety? Using linear discriminant analysis to identify characteristics associated with positive safety rating in London NHS organisations

**DOI:** 10.1101/2021.11.12.21266228

**Authors:** Roberto Fernández Crespo, Ana Luisa Neves, M. Abdulhadi Alagha, Melanie Leis, Kelsey Flott, Owen Bray, Gianluca Fontana, Jess Peck, Vicky Aldred, Ara Darzi

## Abstract

**Objective:** To identify key characteristics associated with a CQC positive and negative safety rating across London NHS organisations.

**Design:** Advanced data analytics and linear discriminant analysis.

**Data sources:** Linked CQC data with patient safety variables sources from 10 publicly available datasets.

**Methods:** Iterative cycles of data extraction, insight generation, and analysis refinement were done and involved regular meetings between the NHS London Patient Safety Leadership Forum and analytic team to optimise academic robustness alongside with translational impact. Ten datasets were selected based on data availability, usability, and relevance and included data from April 2018 to December 2019. Data pre-processing was conducted in R. Missing values were imputed using the median value while empty variables were removed. London NHS organisations were categorised based on their safety rating into two groups: those rated as ‘inadequate’ or ‘requires improvement’ (RI) and those rated as ‘Good’ or ‘outstanding’ (Good). Variable filtering reduced the number of variables from 1104 to 207. The top ten variables with the largest effect sizes associated with Good and RI organisations were selected for inspection. A Linear Discriminant Analysis (LDA) was trained using the 207 variables. Effect sizes and confidence intervals for each variable were calculated. Dunn’s and Kruskal-Wallis tests were used to identify significant differences between RI and Good organisations.

**Results:** Ten variables for Good and RI NHS organisations were identified. Key variables for Good organisations included: Organisation response to address own concerns (answered by nurse/midwife) (Good organisation = 0.691, RI organisation = 0.618, P<.001); fair career progression (answered by medical/dental staff) (Good organisation = 0.905, RI organisation = 0.843, P<.001); existence of annual work appraisal (answered by medical/dental staff)) (Good organisation = 0.922, RI organisation = 0.873, P<.001); organisation’s response to patients’ concerns (Good organisation = 0.791, RI organisation = 0.717, P<.001); harassment, bullying or abuse from staff (answered by AHPHSSP) (Good organisation = 0.527, RI organisation = 0.454, P<.001); adequate materials supplies and equipment (answered by ‘Other’ staff) (Good organisation = 0.663, RI organisation = 0.544, P<.001); organisation response to address own concerns (answered by medical/dental staff) (Good organisation = 0.634, RI organisation = 0.537, P<.001); staff engagement (answered by medical/dental staff) (Good organisation = 0.468, RI organisation = 0.376, P<.001); provision of clear feedback (answered by “other” staff) (Good organisation = 0.719, RI organisation = 0.650, P<.001); and collection of patient feedback (answered by wider healthcare team) (Good organisation = 0.888, RI organisation = 0.804, P<.001).

**Conclusions:** Our study shows that healthcare providers that received positive safety inspections from regulators have significantly different characteristics in terms of staff perceptions of safety than those providers rated as inadequate or requiring improvement. Particularly, organisations rated as good or outstanding are associated with higher levels of organisational safety, staff engagement and capacities to collect and listen to patient experience feedback. This work exemplifies how a partnership between applied healthcare and academic research organisations can be used to address practical considerations in patient safety, resulting in a translational piece of work.

## INTRODUCTION

Over the past two decades, patient safety has undergone an evolution from an emerging domain to a measured indicator and, more recently, to an established cornerstone of healthcare quality, bolstered by robust patient, staff and organisational data.[1] However, the momentum can stall at data capture, leaving meaningful use of data limited.[2] Certainly, patient safety data from incident reporting, staff and patient surveys, organisational audits and many more have been used to drive improvement.

In the UK, the National Reporting and Learning System (NRLS), the national repository of patient safety incidents has been used to issue 150 patient safety alerts from February 2004 to June 2021,[3] thereby mitigating the threat of future incidents. Also in the UK, the “Learning from Litigation Claims” guidance was published in 2021 to support organisations in learning from NHS negligence claims and drive better patient safety.[4] In the US, the analysis of patient complaints has been demonstrated to be an effective way of monitoring diagnostic safety concerns.[5] These examples demonstrate how one data source has yielded singular types of outputs. What is missing is the triangulated use of patient safety data to render more complete and nuanced results. Given that many developed healthcare systems hold millions of data points from diverse sources and given the readily available power of advanced analytic techniques to clean, combine and interrogate these datasets, there is a notable absence of patient safety intelligence driven from multiple data sources. There are many reasons for the deficit in triangulation of patient safety datasets. They are sensitive, not easily shared, inconsistent over time and notoriously incomplete in comparison to other routine healthcare datasets.[6] However, these challenges do not present insurmountable barriers to all prospective data mining, and it is imperative that patient safety datasets are explored alongside each other to gain a holistic picture of safety in any given area. Patient safety improvement is currently compromised due to incomplete and fragmented insights; we could know more, learn more, do more to improve services if our data – and the findings from it - did not remain siloed.

Despite the volume of data and the availability of redacted, publicly available results on patient safety, there is virtually no evidence of a consolidated analysis of them. The NHS Patient Safety Strategy addresses this point and encourages the use of data linkage and advanced analytics techniques to identify emerging issues more quickly.[7] Given the size and scale of the NHS, attempting this required concentrated local efforts before expanding outward to the national level. In January 2020, the NHS London Patient Safety Leadership Forum adopted the challenge set out by the national strategy and led an effort to develop data-driven insights from a range of sources, using advanced analytic techniques. Their ambition centred on the use of publicly available data from acute care providers within London. The integration of patient safety insights can often be compromised because it requires dual expertise: knowledge of the complex safety data landscape as well as the analytics ability to manipulate and draw conclusions from confusing data. The Forum in London addressed this concern by convening a dynamic group of regional NHS policy makers, familiar with patient safety data and clinical priorities as well as a specialist patient safety research centre with dedicated analytic capabilities.

In the UK, the Care Quality Commission (CQC) conducts inspections of health and social care organisations and produces inspection reports that, amongst other considerations, provide a definitive ranking of whether a provider is safe. The CQC safety rating is widely considered the most consistent, reliable, and comprehensive indicator of patient safety in England and uses four ratings: outstanding, good, requires improvement, or inadequate. However, in addition to CQC inspection reports, the English NHS is awash with dozens of other safety data sources, namely NRLS incident reporting, which currently holds more than 20 million records, and patient and staff surveys.[8,9] To date, no study comprehensively triangulated various data sources in patient safety to provide novel insights on the characteristics associated with a positive or negative safety rating. Such analyses could help to understand the practical nuances of what makes a safe organisation, and therefore learn and scale up best practice on patient safety.

### Aim

The main aim of this work was to reveal novel insights on the characteristics associated with a positive safety rating (i.e., CQC safety ratings) in London NHS organisations, using linked datasets and machine learning techniques. Specifically, we triangulated CQC data with patient safety variables sources from 10 publicly available datasets, aiming to identify the variables associated with organisations with a good or outstanding safety rating (Good), and with those rated as inadequate or requiring improvement (RI).

## METHODS

### Description of the datasets

This work used publicly available patient safety data from acute healthcare providers in London. Ten datasets were selected to include in the data mining exercise based on data availability, usability, and relevance (**Table 1**). Data from April 2018 to December 2019 were included.

**Table 1.**
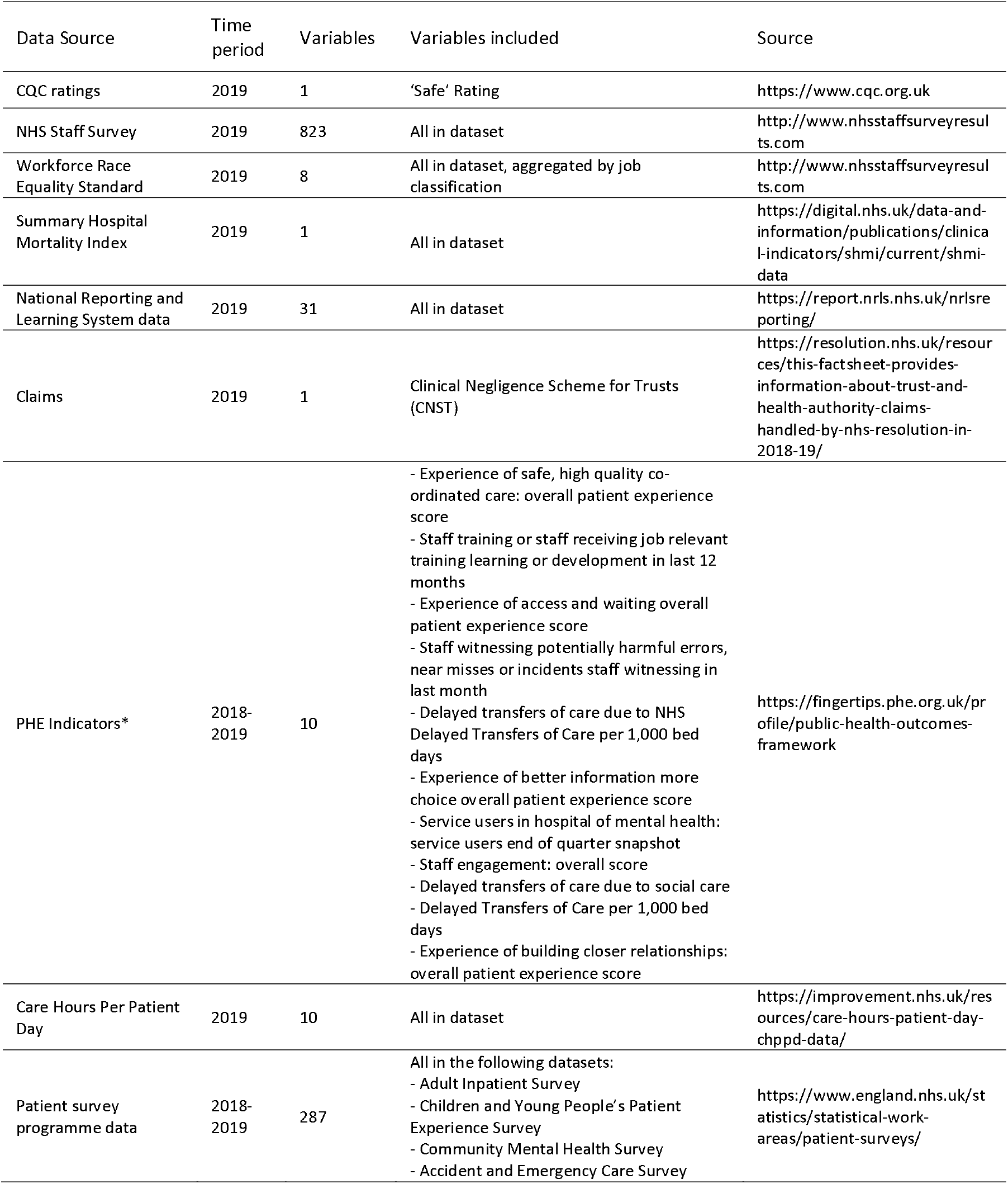
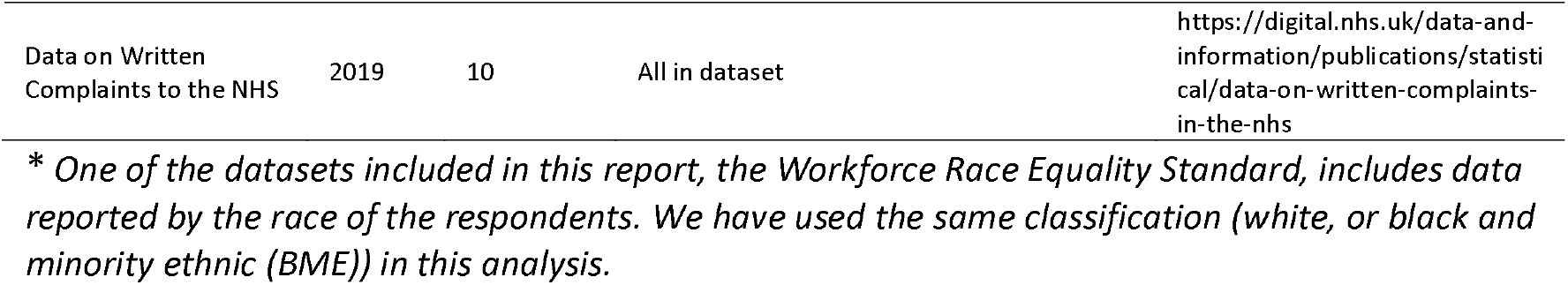
Datasets, time periods, number of variables in the datasets, number of variables included in the final analysis, and sources for each dataset included in the analysis.

| Data Source | Time period | Variables | Variables included | Source |
| --- | --- | --- | --- | --- |
| CQC ratings | 2019 | 1 | 'Safe' Rating | https://www.cqc.org.uk |
| NHS Staff Survey | 2019 | 823 | All in dataset | http://www.nhsstaffsurveyresults.com |
| Workforce Race Equality Standard | 2019 | 8 | All in dataset, aggregated by job classification | http://www.nhsstaffsurveyresults.com |
| Summary Hospital Mortality Index | 2019 | 1 | All in dataset | https://digital.nhs.uk/data-and-information/publications/clinical-indicators/shmi/current/shmi-data |
| National Reporting and Learning System data | 2019 | 31 | All in dataset | https://report.nrls.nhs.uk/nrlsreporting/ |
| Claims | 2019 | 1 | Clinical Negligence Scheme for Trusts (CNST) | https://resolution.nhs.uk/resources/this-fact-sheet-provides-information-about-trust-and-health-authority-claims-handled-by-nhs-resolution-in-2018-19/ |
| PHE Indicators* | 2018-2019 | 10 | - Experience of safe, high quality co-ordinated care: overall patient experience score<br>- Staff training or staff receiving job relevant training learning or development in last 12 months<br>- Experience of access and waiting overall patient experience score<br>- Staff witnessing potentially harmful errors, near misses or incidents staff witnessing in last month<br>- Delayed transfers of care due to NHS Delayed Transfers of Care per 1,000 bed days<br>- Experience of better information more choice overall patient experience score<br>- Service users in hospital of mental health: service users end of quarter snapshot<br>- Staff engagement: overall score<br>- Delayed transfers of care due to social care<br>- Delayed Transfers of Care per 1,000 bed days<br>- Experience of building closer relationships: overall patient experience score | https://fingertips.phe.org.uk/profile/public-health-outcomes-framework |
| Care Hours Per Patient Day | 2019 | 10 | All in dataset | https://improvement.nhs.uk/resources/care-hours-patient-day-chppd-data/ |
| Patient survey programme data | 2018-2019 | 287 | All in the following datasets:<br>- Adult Inpatient Survey<br>- Children and Young People's Patient Experience Survey<br>- Community Mental Health Survey<br>- Accident and Emergency Care Survey | https://www.england.nhs.uk/statistics/statistical-work-areas/patient-surveys/ |
| Data on Written Complaints to the NHS | 2019 | 10 | All in dataset | https://digital.nhs.uk/data-and-information/publications/statistical/data-on-written-complaints-in-the-nhs |

\* One of the datasets included in this report, the Workforce Race Equality Standard, includes data reported by the race of the respondents. We have used the same classification (white, or black and minority ethnic (BME)) in this analysis.

### Data pre-processing

Datasets were imported into R (v. 4.0.0). PHE indicators were accessed and downloaded using the fingertipsR package (v 1.0.3). Only data pertaining to the NHS organisations located in the London area were extracted. Datasets were transformed into a format suitable for analysis using in-house scripts. Empty variables and variables containing identical values for all entries were removed prior to analysis. Missing values in each variable were inputted using the median value.

### Variables

Acute providers in the London region were categorised as ‘outstanding’, ‘good’, ‘requires improvement’, or ‘inadequate’, based on the latest available safety rating as of July 8th, 2020. As of July 8th, 2020, no NHS organisations in London had a rating of ‘outstanding’. The list of organisations, their CQC safety rating, and the classification used for this analysis are shown in **Table 2**. Organisations were categorised into two groups: those that require improvement (RI), which included those that had a safety rating of ‘requires improvement’ or ‘inadequate’, and those that had a safety rating of ‘Good’ (G).

**Table 2.**
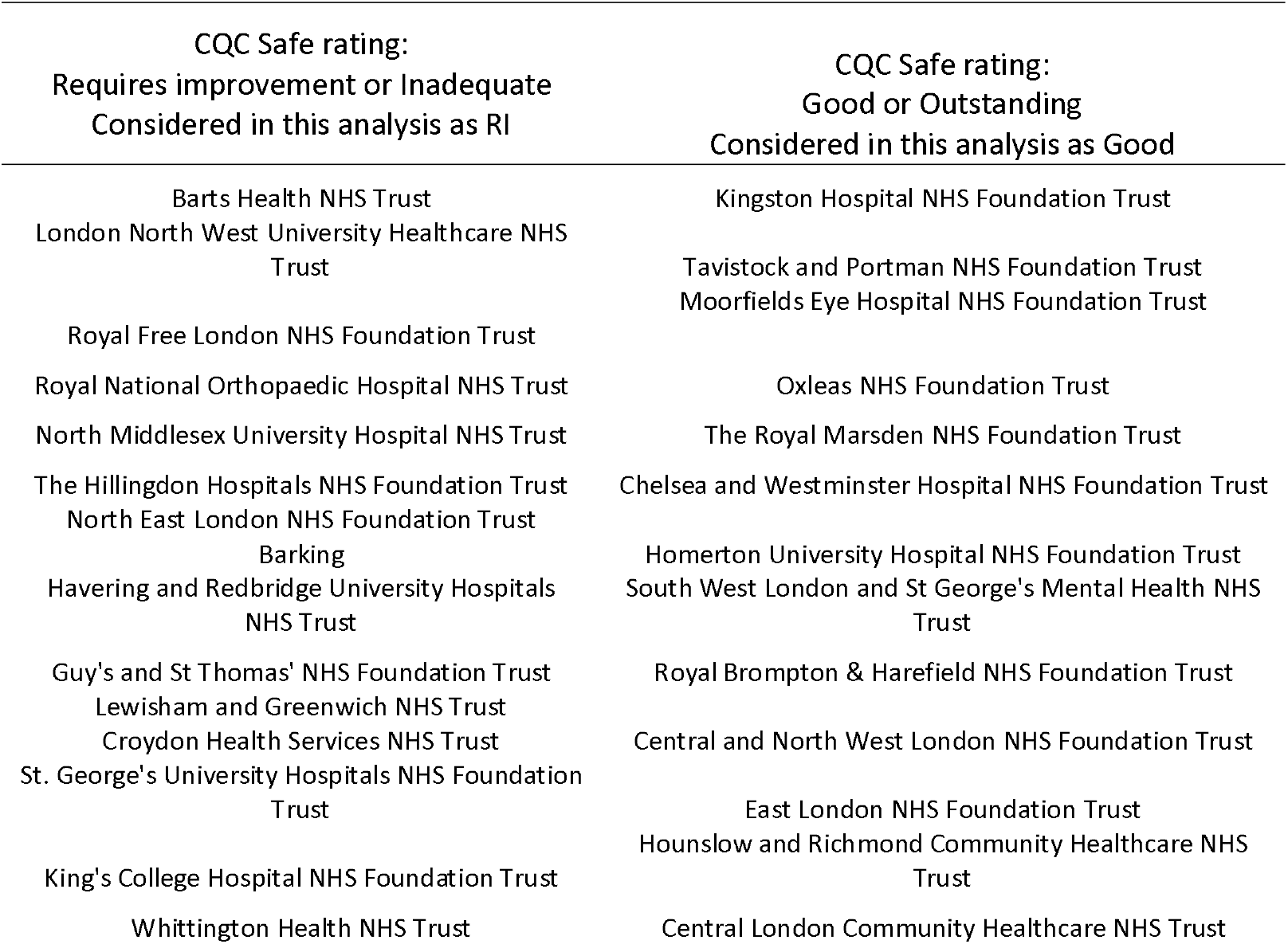
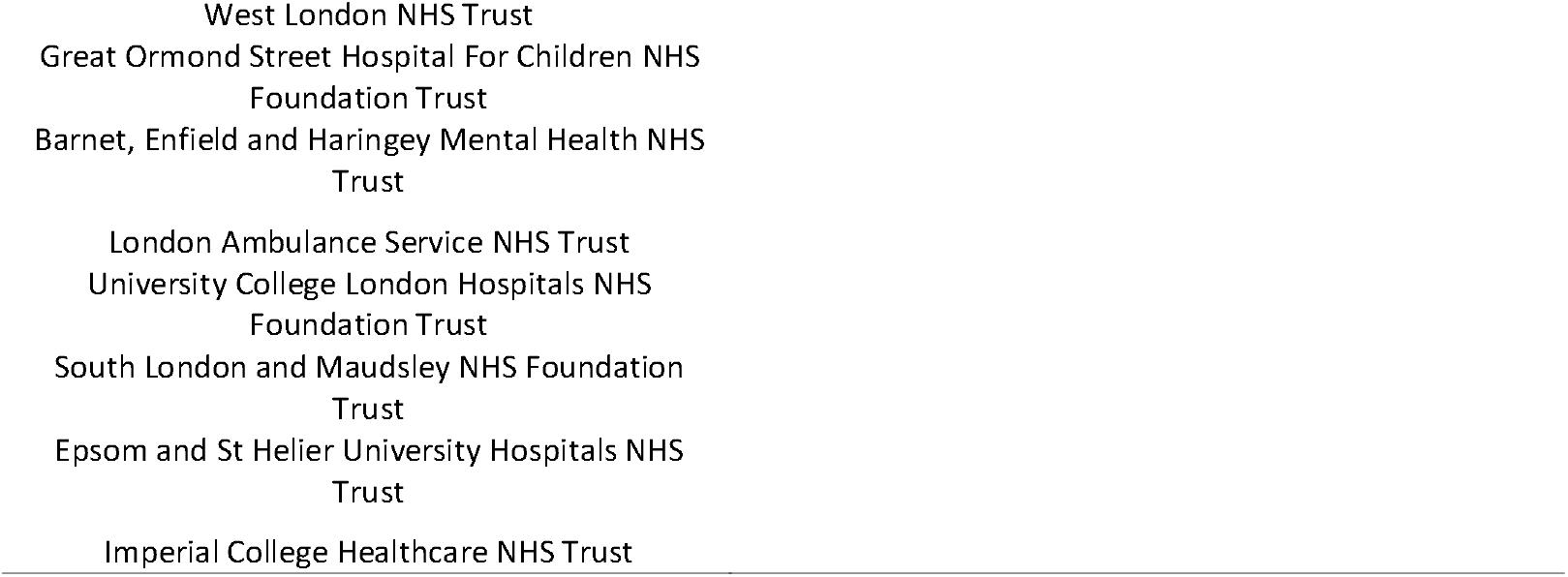
Classification of London NHS Organisations. Care Quality Commission (CQC) ‘Safe’ ratings were extracted on July 8th, 2020.

The initial pool of datasets included 1181 variables. Prior to the analysis, 77 variables were removed due to either having very near-zero variance or not being populated in the original datasets. This led to 1104 variables being used to quantify the contribution of each variable towards the separation of ‘Good’ and ‘RI’ organisations. In order to further identify what variables contributed most significantly towards the separation organisations, the effect size of each variable was also calculated. All variables whose effect size absolute value was over 0.68 were considered for further inspection. This threshold was chosen to allow the selection of variables with sufficiently large effect sizes associated with either ‘Good’ or ‘RI’ organisations, while considering 95% confidence intervals. This filtering step reduced the initial number of variables from 1104 to 207. The number of variables kept for analysis or removed at each stage can be seen in **Figure 1**. The top ten variables with the largest effect sizes associated with ‘Good’ and ‘RI’ organisations were selected for inspection.

**Figure 1.**
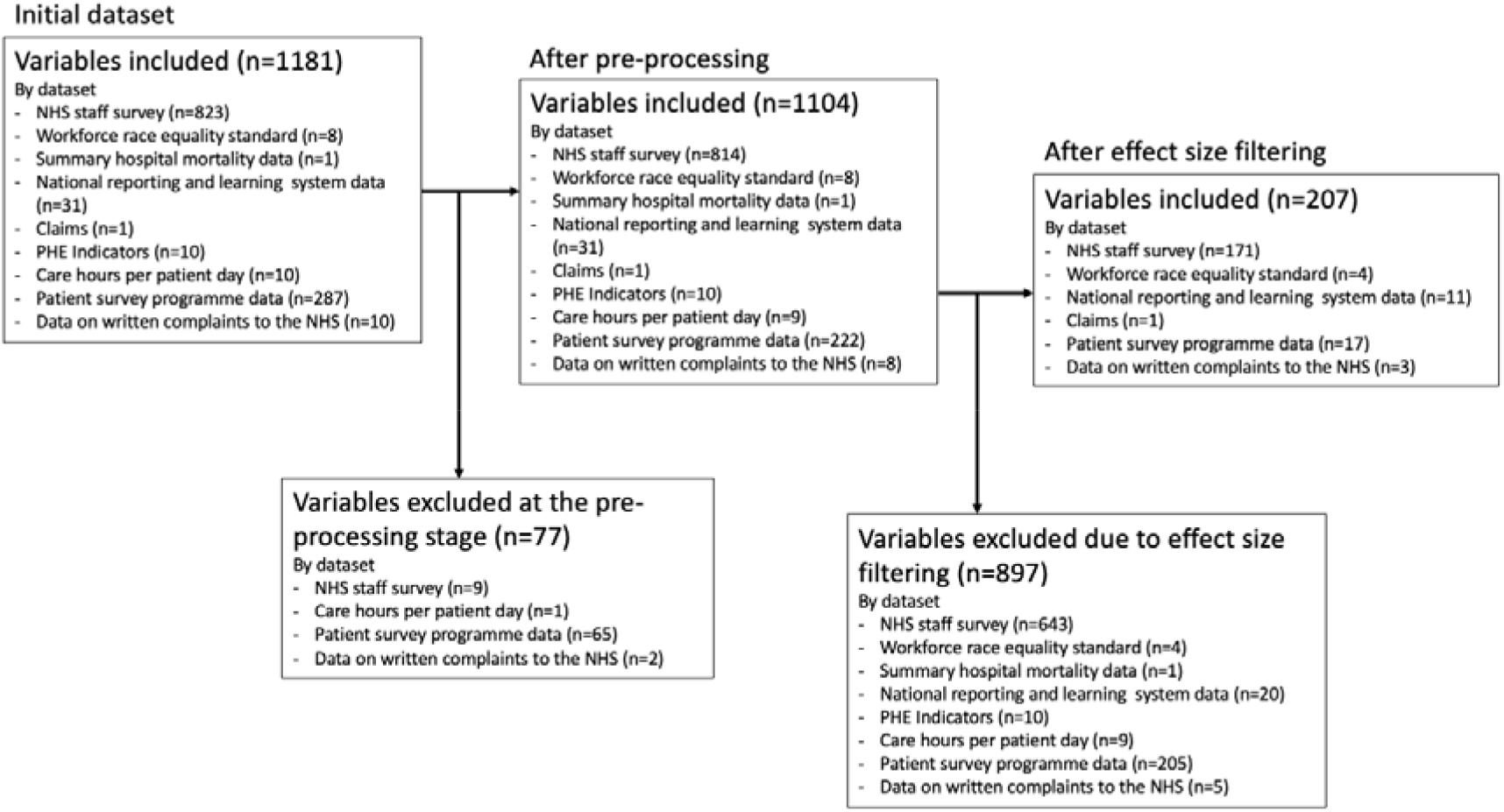
Variable processing workflow.

### Statistical analysis

All statistical analyses were conducted in R (v. 4.0.0). A Linear Discriminant Analysis (LDA) was trained using the variables from the aforementioned datasets, and the contribution of each variable towards the separation of Good and RI organisations was calculated. This was performed using the train function from the caret package (v 6.0-86), specifying the method as ‘lda’. Data were processed prior to modelling by centring the data by subtracting the mean and normalizing it by converting the values to a 0 to 1 scale, removing highly correlated variables, and those with near zero variance. Effect sizes for each variable were calculated using the cohens_d function from the effect size package (v. 0.3.1), also used to calculate 95% confidence intervals for the effect sizes. Dunn’s test was used to identify significant differences in the distribution of individual variables when comparing values for Good and RI organisations. This test was performed using the dunn.test package (v 1.3.5). Significant differences between Good and RI organisations were determined using Kruskal-Wallis test and adjusted for multiple testing using the Benjamin-Höckberg method.

### Stakeholder involvement

This project was carried out through an iterative approach of data extraction, insight generation, and analysis refinement. Regular meetings were held between the NHS London Patient Safety Leadership Forum and analytic team to review findings and refine the analyses to optimise academic robustness alongside with translational impact.

## RESULTS

### Variable contribution and effect size estimation

The contribution of each variable towards the separation of Good and RI organisations was firstly evaluated. The median contribution of each variable was 0.08% (SD = 0.07%). The distribution of the contributions showed a positive skew of 0.617, with 40.03% of the variables having a contribution of 0.01% or less. The top ten contributor variables to the separation between Good and RI organisations can be grouped in five themes: 1) career and feedback (including provision of clear feedback, existence of annual work appraisals, fair career progression, perception that time passes quickly at work); 2) adequate materials supplies and equipment (answered by nurse/midwife, or other staff); 3) patient feedback (including collection of patient feedback and updates on patient feedback); and 4) organisation response (including organisation response to address own concerns (answered by medical/dental, and nurse/midwife), organisation’s response when near missed/incidents are reported errors are reported, and organisation’s response to patients’ concerns) and 5) harassment, bullying or abuse from staff. A full description of the ten contributor variables is provided in **Table 3**.

**Table 3.** Top ten variables, including ties, that contributed the most towards the separation of Good and RI organisations

| Variable | Question | Source | Rank | Effect size |
| --- | --- | --- | --- | --- |
| Provision of clear feedback (a) | My immediate manager, who may be referred to as your line manager, gives me clear feedback on my work (a) | NHS Staff survey | 1 | 1.25 (0.525-1.958) |
| Fair career progression (b) | Does your organisation act fairly with regard to career progression/ promotion regardless of ethnic background, gender, religion, sexual orientation, disability or age (b) | NHS Staff survey | 2 | 1.434 (0.689-2.161) |
| Organisation response to address own concerns (c) | I am confident that my organisation would address my concern (c) | NHS Staff survey | 3 | 1.491 (0.739-2.225) |
| Harassment, bullying or abuse from staff (d) | Harassment bullying or abuse from staff in last 12 months (d) | NHS Workforce Race Equality Standard Survey | 4 | -1.293 (-2.006 - -0.564) |
| Existence of annual work appraisal (b) | In the last 12 months have you had an appraisal, annual review, development review, or Knowledge and Skills Framework KSF development review (b) | NHS Staff survey | 5 | 1.374 (0.636-2.095) |
| Adequate materials supplies and equipment (c) | I have adequate materials supplies and equipment to do my work (c) | NHS Staff survey | 6 | 1.138 (0.425-1.836) |
| Adequate materials supplies and equipment (a) | I have adequate materials supplies and equipment to do my work (a) | NHS Staff survey | 6 | 1.295 (0.566-2.008) |
| Updates on patient feedback (b) | I receive regular updates on patient service user experience feedback in my directorate department e.g., via line managers or communications teams (b) | NHS Staff survey | 8 | 1.156 (0.441-1.855) |
| Collection of patient feedback collected (e) | Is patient service user experience feedback collected within your directorate department (e.g., Friends and Family Test patient surveys etc) (e) | NHS Staff survey | 10 | 1.246 (0.522-1.954) |
| Perception that time passes quickly (f) | Time passes quickly when I am working (f) | NHS Staff survey | 10 | 1.107 (0.397-1.802) |
| Organisation's response to patients' concerns (f) | My organisation acts on concerns raised by patients, service users (f) | NHS Staff survey | 10 | 1.325 (0.592-2.041) |
| Organisation's response when near missed/incidents are reported errors are reported (b) | When errors near misses or incidents are reported my organisation takes action to ensure that they do not happen again (b) | NHS Staff survey | 10 | 1.158 (0.443-1.858) |
| Organisation response to address own concerns (b) | I am confident that my organisation would address my concern (b) | NHS Staff survey | 10 | 1.294 (0.565-2.007) |

(a) answered by "other" staff; (b) answered by medical/dental staff; (c) answered by nurse/midwife; (d) answered by white staff; (e) answered by wider healthcare team; (f) answered by allied health professionals, healthcare scientists or scientific technical staff.

All top ten contributor variables appeared in the NHS Staff survey questionnaire, except for one variable which belonged to the Workforce Race Equality Standard Survey (i.e., harassment, bullying or abuse from staff in the last 12 months, answered by white staff).

### Variables associated with Good or RI organisations

The top ten variables with the largest effect sizes associated with Good and RI organisations were selected for inspection (**Figure 2A, Table 4**). For NHS organisations classified as Good (G), ten variables were identified: Organisation response to address own concerns (answered by nurse/midwife) (Good organisation = 0.691, RI organisation = 0.618, P<.001); fair career progression (answered by medical/dental staff) (Good organisation = 0.905, RI organisation = 0.843, P<.001); existence of annual work appraisal (answered by medical/dental staff)) (Good organisation = 0.922, RI organisation = 0.873, P<.001); organisation’s response to patients’ concerns (Good organisation = 0.791, RI organisation = 0.717, P<.001); harassment, bullying or abuse from staff ((answered by AHPHSSP) (Good organisation = 0.527, RI organisation = 0.454, P<.001); adequate materials supplies and equipment (answered by ‘Other’ staff) (Good organisation = 0.663, RI organisation = 0.544, P<.001); organisation response to address own concerns (answered by medical/dental staff) (Good organisation = 0.634, RI organisation = 0.537, P<.001); staff engagement (answered by medical/dental staff) (Good organisation = 0.468, RI organisation = 0.376, P<.001); provision of clear feedback (answered by “other” staff) (Good organisation = 0.719, RI organisation = 0.650, P<.001); and collection of patient feedback (answered by wider healthcare team) (Good organisation = 0.888, RI organisation = 0.804, P<.001). These variables had an overarching theme of NHS workers’ having a positive experience at work and all of them were sourced from the NHS Staff survey. An overview of the differences between Good and RI organisations, for each variable, is provided in **Figure 2**.

**Figure 2.**
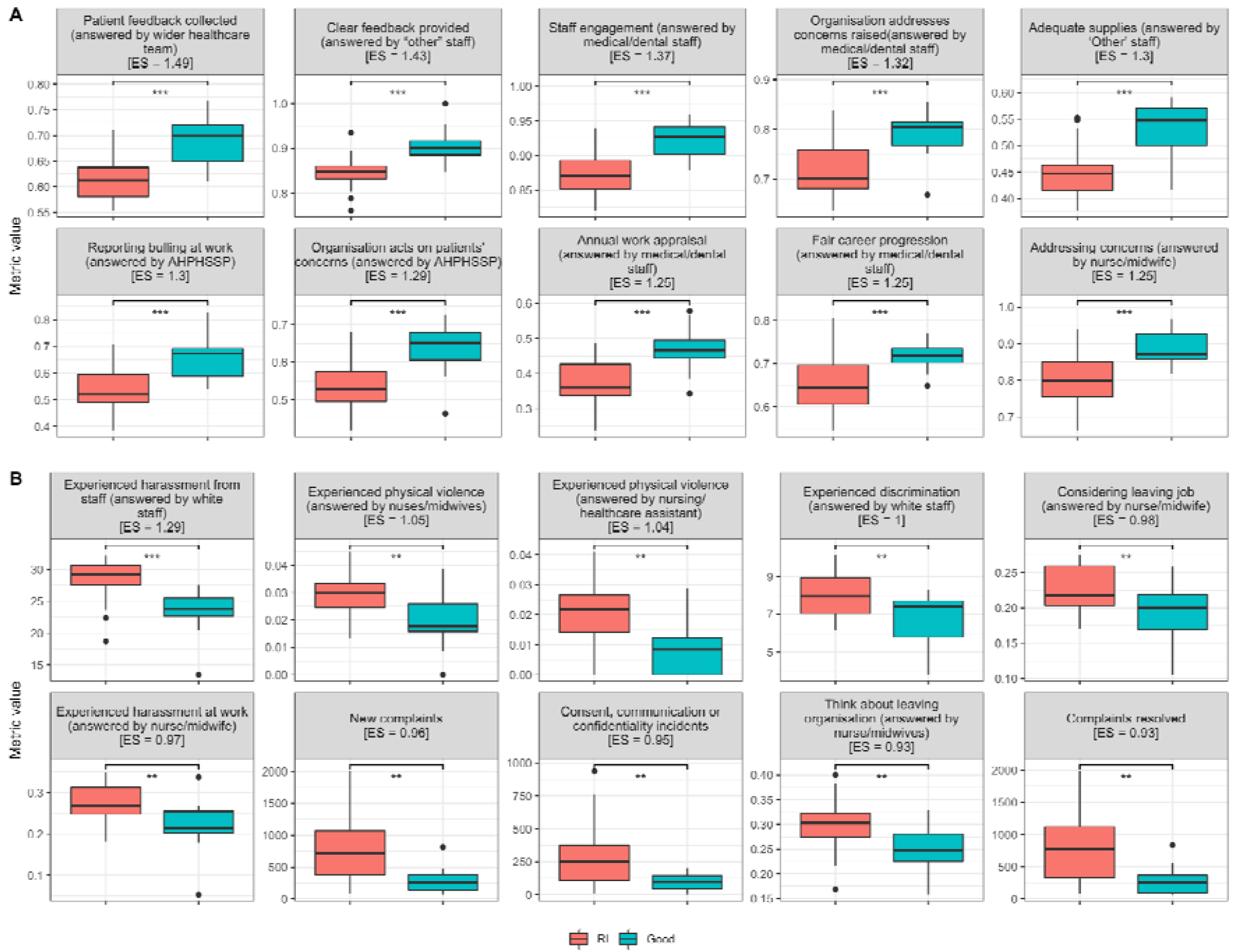
Top ten variables with the largest effect size (ES) for organisations classified as Good (A), and RI (B). For each variable, a boxplot showing the distribution of the values for RI organisations (shown in red) and Good organisations (shown in blue) is displayed. The banners on top of the plots indicate the variable shown, plus the ES for this variable (in brackets). Staff survey responses are measured as average question value (values closer to one being more positive answers), complaints (new and resolved) as the total number of complaints received within the year, and incidents received within a year.

**Table 4.**
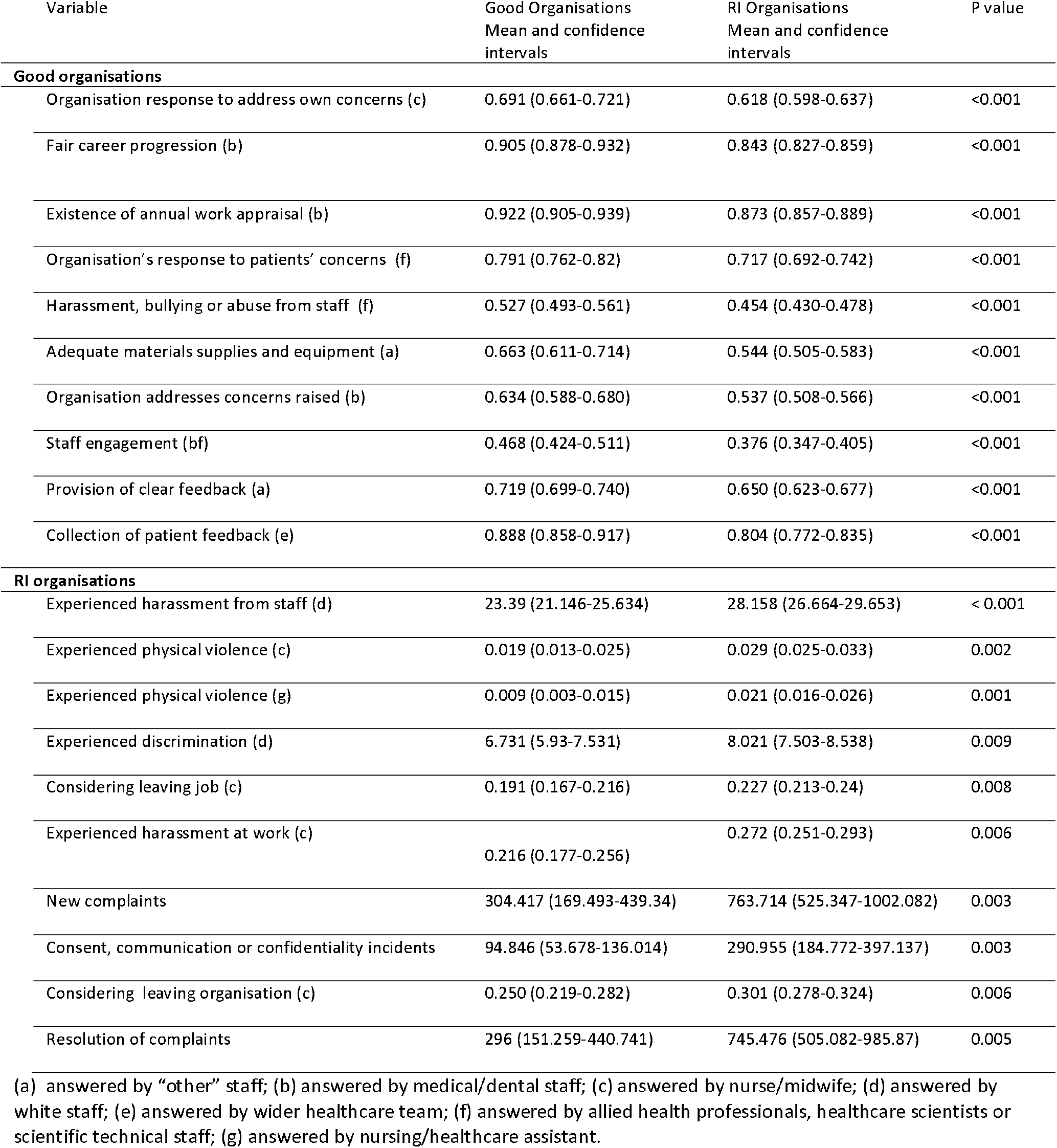
Top ten variables associated with Good and RI organisations.

| Variable | Good Organisations<br>Mean and confidence<br>intervals | RI Organisations<br>Mean and confidence<br>intervals | P value |
| --- | --- | --- | --- |
| **Good organisations** |  |  |  |
| Organisation response to address own concerns (c) | 0.691 (0.661-0.721) | 0.618 (0.598-0.637) | <0.001 |
| Fair career progression (b) | 0.905 (0.878-0.932) | 0.843 (0.827-0.859) | <0.001 |
| Existence of annual work appraisal (b) | 0.922 (0.905-0.939) | 0.873 (0.857-0.889) | <0.001 |
| Organisation's response to patients' concerns (f) | 0.791 (0.762-0.82) | 0.717 (0.692-0.742) | <0.001 |
| Harassment, bullying or abuse from staff (f) | 0.527 (0.493-0.561) | 0.454 (0.430-0.478) | <0.001 |
| Adequate materials supplies and equipment (a) | 0.663 (0.611-0.714) | 0.544 (0.505-0.583) | <0.001 |
| Organisation addresses concerns raised (b) | 0.634 (0.588-0.680) | 0.537 (0.508-0.566) | <0.001 |
| Staff engagement (bf) | 0.468 (0.424-0.511) | 0.376 (0.347-0.405) | <0.001 |
| Provision of clear feedback (a) | 0.719 (0.699-0.740) | 0.650 (0.623-0.677) | <0.001 |
| Collection of patient feedback (e) | 0.888 (0.858-0.917) | 0.804 (0.772-0.835) | <0.001 |
| **RI organisations** |  |  |  |
| Experienced harassment from staff (d) | 23.39 (21.146-25.634) | 28.158 (26.664-29.653) | < 0.001 |
| Experienced physical violence (c) | 0.019 (0.013-0.025) | 0.029 (0.025-0.033) | 0.002 |
| Experienced physical violence (g) | 0.009 (0.003-0.015) | 0.021 (0.016-0.026) | 0.001 |
| Experienced discrimination (d) | 6.731 (5.93-7.531) | 8.021 (7.503-8.538) | 0.009 |
| Considering leaving job (c) | 0.191 (0.167-0.216) | 0.227 (0.213-0.24) | 0.008 |
| Experienced harassment at work (c) | 0.216 (0.177-0.256) | 0.272 (0.251-0.293) | 0.006 |
| New complaints | 304.417 (169.493-439.34) | 763.714 (525.347-1002.082) | 0.003 |
| Consent, communication or confidentiality incidents | 94.846 (53.678-136.014) | 290.955 (184.772-397.137) | 0.003 |
| Considering leaving organisation (c) | 0.250 (0.219-0.282) | 0.301 (0.278-0.324) | 0.006 |
| Resolution of complaints | 296 (151.259-440.741) | 745.476 (505.082-985.87) | 0.005 |

(a) answered by "other" staff; (b) answered by medical/dental staff; (c) answered by nurse/midwife; (d) answered by white staff; (e) answered by wider healthcare team; (f) answered by allied health professionals, healthcare scientists or scientific technical staff; (g) answered by nursing/healthcare assistant.

The top ten variables with the largest effect size associated with RI organisations included variables from the NHS Staff Survey, the complaints dataset, NRLS and the NHS workforce race equality survey (**Figure 2B, Table 4**). For NHS organisations classified as RI, ten identified variables were: Experienced harassment from staff (answered by white staff) (Good organisation = 23.390, RI organisation = 28,158 ,p. value < 0.001); Experienced physical violence (answered by nurses/midwives) Good organisation = 0.019, RI organisation = 0.029, p. value = 0.002); Experienced physical violence (answered by nursing/healthcare assistant) (Good organisation = 0.009, RI organisation = 0.021 ,p. value = 0.001); Experienced discrimination (answered by white staff) (Good organisation = 6.731, RI organisation = 8.02 1,p. value = 0.009); Considering leaving job (answered by nurse/midwife) (Good organisation = 0.191, RI organisation = 0.227 ,p. value = 0.008); Experienced harassment at work (answered by nurse/midwife) (Good organisation = 0.216, RI organisation = 0.272,p. value = 0.006); New complaints (Good organisation = 304.417, RI organisation = 763.714,p. value = 0.003); Consent, communication or confidentiality incidents (Good organisation = 94.846, RI organisation = 290.955, p. value = 0.003); Think about leaving organisation (answered by nurse/midwives) (Good organisation = 0.250, RI organisation = 0.301 ,p. value = 0.006); Complaints resolved (Good organisation = 296.00, RI organisation = 754.476,p. value = 0.005). The variables were generally related to NHS workers’ having a negative experience at work.

## DISCUSSION

### Key findings

This study triangulated ten routinely collected London NHS datasets and sought to underpin some of the key factors associated with safer hospitals using linear discriminant analysis. Good Organisations were defined as those with CQC Safe rating of “Good” or “Outstanding” while RI Organisations encompassed the rest of the rating scale; “Inadequate” or “Requires improvement”. Our findings suggest that the enhancement of safety rating primarily requires the deliberate recognition and management of three stakeholders’ needs; staff, patients, and organisations (as inferred from NHS Staff Survey).

Good Organisations are positively associated with aspects related to staff engagement (including fair career progression, existence of annual appraisal, provision of clear feedback, staff engagement, harassment incident reporting); listening to patients’ voice; and organisational factors (including adequate materials supplies and equipment as well as the organisations’ ability to address both the individual’s and patients concerns). In contrast, RI Organisations have a higher number of adverse incidents including complaints; communication; confidentiality; harassment; discrimination and physical violence.

### Comparison with previous literature

The aviation industry developed a framework [10], based on Reason’s Swiss Cheese model, for evaluating adverse events through a human factors lens (Human Factors Analysis and Classification System - HFACS) and was also adapted to promote patient and workforce safety.[11] Central to understanding the challenges facing organisation’s safety is the explicit articulation of the needs of each party.

#### Staff Engagement

Staff who develop a sense of attachment and investment towards their organisation were shown to perform 20% better than their colleagues.[12] There is also compelling evidence in healthcare of the association between staff engagement and quality of care and patient safety outcomes.[13,14] In line with previously conceptualised staff engagement frameworks,[15] our study highlights the importance of addressing staff expectations through a) fair career progression opportunities, b/c) the provision of meaningful feedback and annual appraisals and d) building capacities that value and encourage harassment reporting. In other words, healthcare workers self-reported perceptions of how they receive feedback, as well as their perceptions of fair treatment regardless of personal characteristics, were influential determinants of organisational safety.

#### Listening and Acting on Patients’ Voice

Patient experience feedback data is routinely collected in the NHS for various purposes, but less effort, clarity and consistency go into analysing it to drive effective improvements. The National Institute for Health Research (NIHR) [16] compiled the findings of recent studies on the use of NHS patient experience data and found that the majority of analyses are presented to managerial- and corporate-level teams, rather than engaging frontline staff, local units and patients who are likely to co-design interventions that lead to sustainable improvements.[17] Our study shows that Good Organisations appear to incorporate feedback from patients and users more than RI Organisations. However, future studies are required to develop effective evidence-based policies that act on patient feedback at both local and national levels.

#### Organisational Safety

With regards to Reason’s model [18] and HFACS framework, our findings re-draw the attention that issues related to organisational influences and senior management are latent errors and have a direct impact on the other levels of human failure. These failures encompass (ref) three domains: resource management, organisational processes, and organisational climate. For Organisations to be “Good” and efficient, safety professionals should develop a safety culture within organisations by having a robust incident reporting of errors and near misses (*organisational climate*),[19,20] ensuring adequate and timely materials and equipment supplies (*resource management*) and co-design and practise evidence-based protocols that addresses individual and patients concerns (*organisational process*).[21]

### Strengths and limitations

Our study is the first of its kind to triangulate ten routinely collected patient safety datasets across London NHS Organisations and uses advanced analytics to derive patient safety insights. It highlights the potential of advanced analytics to uncover novel insights in the field of patient safety using publicly available data routinely collected healthcare data and cements the importance of using multiple sources of intelligence to derive patient safety insights, as suggested by the NHS Patient Safety Strategy.[7]

A unique feature of this work in fact was the partnership between applied healthcare and academic research organisations that ensured that the academics’ technical expertise was used to address practical considerations in patient safety in London, resulting in a truly translational piece of work. Outputs from this work have been shared broadly across the NHS, with this co-production model serving as a blueprint for future collaboration on translational projects.

The LDA model used in conjunction with effect size estimation provided a method of identifying what variables contributed the most towards the separation of the two groups of organisations. Calculating the effect size of each variable provides a way to identify those variables that could characterize each of the two groups of organisations. Combined with the variable contributions from the LDA model, the data sources that could be the most relevant towards characterising these two groups were identified. Adding on to the results seen by only LDA contributions, the NHS Staff Survey and NHS Workforce Race Equality data were the two predominant datasets. While this could be expected of the staff survey given the large number of variables included in this dataset, 50% of the variables from the race equality survey were selected after effect size filtering, possibly indicating the importance of this dataset relative to the number of variables included in it.

Some limitations must also be acknowledged. Although this study used extensive data from diverse sources, the team identified others that could have been relevant, but were either not publicly available or not easily accessible in a machine-readable format (i.e., Never Events data,[22] Hospital Episode Statistics (HES) / Office for National Statistics (ONS) linked mortality data.[23] These options should all be considered in future work as they are valuable sources of patient safety-related data. For example, data from the Safety Thermometer survey, among other staff and patient feedback data, have been used in the past to try to predict safety outcomes and find correlates to perceived safety and safety outcomes.[1]

Furthermore, the time frame of each dataset should also be considered. While each of the datasets included here are the latest available (as of July 8^th^, 2020), datasets such as PHE Fingertips are published on a quarterly or monthly basis, depending on the variable. The results from the NHS Staff Survey are published yearly, while not all Patient Surveys are published every year. Another important consideration is the safety classification of healthcare providers. For this analysis, we have utilised the CQC safety rating. Future analysis could explore the possibility of a more nuanced classification of organisations that may capture other variables.

### Implications for policy and research

Patient safety is an established and broadly recognised pillar of healthcare quality, and this recognition has supported the collection of extensive data related to it. It has provoked the movement to regulate healthcare safety standards and assess providers along these parameters. The progress in collecting this data and creating local, national, even international, databases of patient safety data is an important achievement. However, the volume of this data demands more advanced use. It is becoming less justifiable to examine patient safety according to individual data points, and we are increasingly realising the ethical implications of capturing so much data and not optimising its use.

From a research perspective, it is also important to further evaluate the differences observed in the responses from diverse groups of persons. In what concerns the question on harassment, bullying or abuse from staff, the question answered by white staff ranked fourth in our list of top variables, while the one answered by BME staff ranked 237th out of 1104. For all four questions included in the race equality survey, answers by white staff were ranked higher in terms on LDA contribution. Future research could be conducted to explain these differences.

Equally, the question evaluating material supplies appeared in the list of top variables as answered by registered nurses/midwives and individuals included in the ‘Other’ category, but the impact for other staff groups may be different. Future research should focus on explaining the differences across working groups when answering this set of questions.

### Conclusion

Our results show that healthcare providers that received positive safety inspections from regulators have significantly different characteristics in terms of staff perceptions of safety than those providers rated as requiring improvement. This triangulated use of data not only allows more comprehensive insights about organisations, but it also gives weight to existing safety ratings. As health systems continue to capture more patient safety data, it is increasingly important that this data is used and that patient safety decisions are made in consideration of multiple sources of intelligence.

## Data Availability

All data produced are available online at their respective sources (see Table 1).

## ACKNOWLEDGEMENTS

None

## ETHICAL APPROVAL

Was not required for this project given that the data used is publicly available.

